# Clinical genetics lacks standard definitions and protocols for the collection and use of diversity measures

**DOI:** 10.1101/2020.04.22.20074500

**Authors:** Alice B. Popejoy, Kristy R. Crooks, Stephanie M. Fullerton, Lucia A. Hindorff, Gillian W. Hooker, Barbara A. Koenig, Natalie Pino, Erin M. Ramos, Deborah I. Ritter, Hannah Wand, Matt W. Wright, Michael Yudell, James Y. Zou, Sharon E. Plon, Carlos D. Bustamante, Kelly E. Ormond, on behalf of the Clinical Genome Resource (ClinGen) Ancestry and Diversity Working Group (ADWG)

**Author notes:** These authors contributed equally (Carlos D. Bustamante, Kelly E. Ormond).

## Abstract

Genetics researchers and clinical professionals rely on diversity measures such as race, ethnicity, and ancestry (REA) to stratify study participants and patients for a variety of applications in research and precision medicine. However, there are no comprehensive, widely accepted standards or guidelines for collecting and using such data in either setting. Two NIH-funded research consortia, the Clinical Genome Resource (ClinGen) and Clinical Sequencing Evidence-generating Research (CSER), have partnered to address this issue and report how REA are currently collected, conceptualized, and used. Surveying clinical genetics professionals and researchers (N=448), we found heterogeneity in the way REA are perceived, defined, and measured, with variation in the perceived importance of REA in both clinical and research settings. The majority of respondents (>55%) felt that REA are at least somewhat important for clinical variant interpretation, ordering genetic tests, and communicating results to patients. However, there was no consensus on the relevance of REA, including how each of these measures should be used in different scenarios and what information they can convey in the context of human genetics. A lack of common definitions and applications of REA across the precision medicine pipeline may contribute to inconsistencies in data collection, missing or inaccurate classifications, and misleading or inconclusive results. Thus, our findings support the need for standardization and harmonization of REA data collection and use in clinical genetics and precision health research.

## Introduction

Different aspects of human diversity are captured by the concepts of “race,” “ethnicity”, and “ancestry” (REA). While these terms are widely used in biomedical research, clinical care, and reporting health statistics for federal and state funding in the United States (NOT-OD-15-089), they are not interchangeable, and they are important to define. While there are no universal definitions, we consider “race” and “ethnicity” to be socio-cultural factors that provide information about environmental exposures but are not directly indicative of genetic risk factors for disease.

The related but distinct concept of “ancestry,” referring to the genetic inheritance of variants from global ancestral populations, has increasingly been used in genomics research and has implications for certain clinical applications of genetics such as assessing whether a variant is rare or common in a particular ancestral population (Kumar et al., 2010). Ancestry as a genetic concept has also gained prominence in the public sphere due to the rise in popularity of direct-to-consumer genetic testing products (Royal et al., 2010; Jorde & Bamshad, 2020). However, there are no standard definitions of these terms in medical genetics and no consensus on protocols for the collection or estimation of this information across research institutions or healthcare systems. For example, our prior work demonstrated that the categorization of diversity measures on clinical requisition forms varies widely among genetic testing laboratories (Popejoy et al., 2018). Importantly, it is unclear to what extent this information is used to interpret genetic test results and whether inconsistent data collection among laboratories contributes to variation in reporting and delivery of clinical genetics, including variant classification, application of screening guidelines, risk elimination, or other measures relevant to clinical care.

Previous studies have examined how race is conceptualized by physicians and genetics professionals (Bonham et al., 2009; Bonham et al., 2014; Nelson et al., 2019). The ways in which REA influence health outcomes, particularly as they inform health disparities and population-based treatment or interventions, is also an active area of research (Cunningham et al., 2015). The literature has shown that definitions of “race and “ethnicity” are historically fluid, context-specific, and often used interchangeably (Race, Ethnicity, and Genetics Working Group, 2005; Smedley & Smedley, 2012; Omi & Winant, 2014; Roth, 2016). The conflation of these concepts, as well as limitations in the type of information available in different settings, also leads to “race” and/or “ethnicity” being used as a proxy for genetic ancestry or genomic background. This practice may disproportionately disadvantage people from diverse racial, ethnic, and ancestral backgrounds because self-reported identity measures are based on incomplete knowledge, are often inaccurate representations of ancestry, and those with multiple ancestries are not easily grouped into discrete racial or ethnic categories (Braun et al., 2007). Furthermore, the lack of diversity in genomic databases means that even if self-reported measures serve as a reasonable proxy for ancestral background, this information may have limited utility and actionability in clinical genetics (Landry et al., 2018).

The Ancestry and Diversity Working Group (ADWG) of the NIH-funded Clinical Genome Resource (ClinGen, http://clinicalgenome.org) is a multi-disciplinary group of investigators from academic research institutions and hospitals across the United States. ClinGen is an important knowledgebase for clinical genetics, including a set of FDA-recognized variant-disease expert curations. The ClinGen ADWG is responsible for ensuring that this resource is representative of and responsive to a broad spectrum of human diversity. ADWG members have expertise spanning genomics, epidemiology, public health, bioethics, and social sciences. Positioned at the intersections of these diverse professional perspectives, the ADWG is intended to conduct research and provide evidence-based guidance about the use of REA in clinical genetics.

Another NIH-funded research consortium, Clinical Sequencing Evidence-generating Research (CSER, https://cser-consortium.org), is a collaborative effort of seven independent research sites conducting studies in clinical translational genetics with a specific focus on diverse populations. CSER investigators are also working to address disparities in clinical genetics and the development of standards for the use of REA. This study is the result of a collaborative effort between the two consortia to determine how REA are conceptualized, utilized, and communicated in clinical genetics research and practice.

While there may be some effective and necessary uses of REA data, such as contextualizing the genomic or environmental background of a genetic variant with potential clinical significance, continued collection and use of these measures without a clear justification and framework may lead to differential clinical treatment and quality of care among racial and ethnic minority groups with negative implications more broadly for the fair distribution of benefits from precision medicine research. Despite the tradeoffs between the potential utility and harms of racial and ethnic classification (Mays et al., 2002), no study to date has conducted a comprehensive assessment of how the terms “race”, “ethnicity”, and “ancestry” are understood by clinical genetics professionals and researchers, or how these perceptions inform clinical genetic testing, variant interpretation, reporting, diagnosis, and treatment. The survey described in this paper represents the first investigation of its kind, interrogating both perceptions and reported use of REA among clinical genetics professionals.

## Subjects & Methods

The study included both non-clinical genetics researchers and clinical genetics professionals (i.e., clinical geneticists, genetic counselors, clinical laboratory directors, and other clinical laboratory employees). Most questions were designed for clinical genetics professionals and related to clinical activities such as test ordering, variant interpretation, and reporting genetic test results to patients.

In total, 121 survey questions (see Appendix) were developed and refined through an iterative process by ClinGen ADWG and CSER investigators. An initial set of survey questions was developed and approved by ADWG members, reviewed and revised based on feedback from CSER investigators, and further revised based on feedback from 11 cognitive interviews of individuals representative of those being surveyed (Supplemental methods contain details about how these interviews were conducted).

The survey included required multiple-choice questions about respondents’ perceived definitions and utility of REA in research and clinical genetics; these questions were developed based on responses to a previous study of physician perspectives on race and ethnicity (Bonham et al., 2014). Optional participant demographic questions included inquiries about personal identity: 1) free-text boxes for “sex”, “gender”, “race”, “ethnicity”, and “ancestry” and 2) multiple-choice options reflecting the categories provided on the U.S. Census and approved by the U.S. Office of Management and Budget (OMB). Self-reported professional demographics (career type, professional experience, and frequency of conducting variant classification for clinical or research purposes, ordering genetic tests and reporting results to patients) were used to create branching logic to ensure the relevance of each question to each survey respondent. The remaining questions covered the type and source(s) of REA data used in clinical gene and variant curation and interpretation, clinical contextual factors impacting the interpretation and communication of genetic testing results to patients, and opinions on whether guidelines are needed for the clinical genetics community around the use of such diversity measures. These questions included a combination of multiple-choice, true-false, and free-text response options.

### Institutional Review

The Stanford University Institutional Review Board (IRB) determined the proposed study did not meet the definition of research or clinical investigation on human subjects and was therefore exempt from IRB review.

### Data Collection

The online survey was administered from September 2018 to April 2019 through Stanford Qualtrics with separate recruitment links for each disseminating organization: ClinGen (N=788), CSER (N=184), the American Board of Genetic Counselors (ABGC, N=4661), the American Society of Human Genetics (ASHG, N=2659), and the American College of Medical Genetics and Genomics (ACMG, N=2218). A follow-up recruitment email was also sent to the ClinGen and CSER listservs, as well as a manually curated subset of clinical geneticists in the United States whose emails are publicly available (N=638). Table S1 shows the number of individuals contacted through each organization and their corresponding response rate estimates. After response rates were estimated for each disseminating organization and follow-up emails were sent to the initial solicitations, a snowball approach was adopted in February and March 2019. The purpose of the snowball approach was to reach additional target participants through individual emails to colleagues and professional networks, and social media such as public posts on Facebook and Twitter. There was no incentive provided for completing the survey.

### Data Analysis

Data were downloaded from Qualtrics and analyzed using a custom Python script. The majority of questions were designed to assess current practices in research and clinical genetics and as such most data analysis involved descriptive statistics. For certain analyses, responses on a Likert scale were combined to show broad categories of agreement (e.g., “well” / “very well” and “often” / “always”). The majority of findings showed no trends in response themes between different types of participants (e.g., clinical professionals vs. non-clinical researchers, most experienced vs. least experienced professionals, etc.).

## Results

### Study Participants

A total of 448 respondents completed the survey, including non-clinical researchers (N=87), clinical genetics professionals (N=268), trainees (N=12) and others. For all types of respondents, we report on demographics, perceived definitions of REA, and opinions on the need for guidelines. In order to gain a sense of how REA are used in clinical genetics, we focus on results obtained from clinical genetics professionals, identified by their reported professional roles and activities.

Table 1 indicates the primary employment affiliations, professional roles, years of experience, and clinical expertise of respondents. Most respondents were clinical genetics professionals who are employed at academic or commercial diagnostic institutions in the United States. Of the respondents who provided information about their professional experience, 55% (N=218) had 1-10 years of professional experience, 24% (N=96) had between 11-20 years of experience, and 21% (N=84) had >20 years. Among those who provided information about their professional roles (N=398), 42% were genetic counselors (N=168); others included clinical geneticists (N=57, 14%), clinical laboratory directors (N=43, 11%), non-clinical researchers (N=87, 22%), and trainees (N=12, 3%).

**Table 1.**
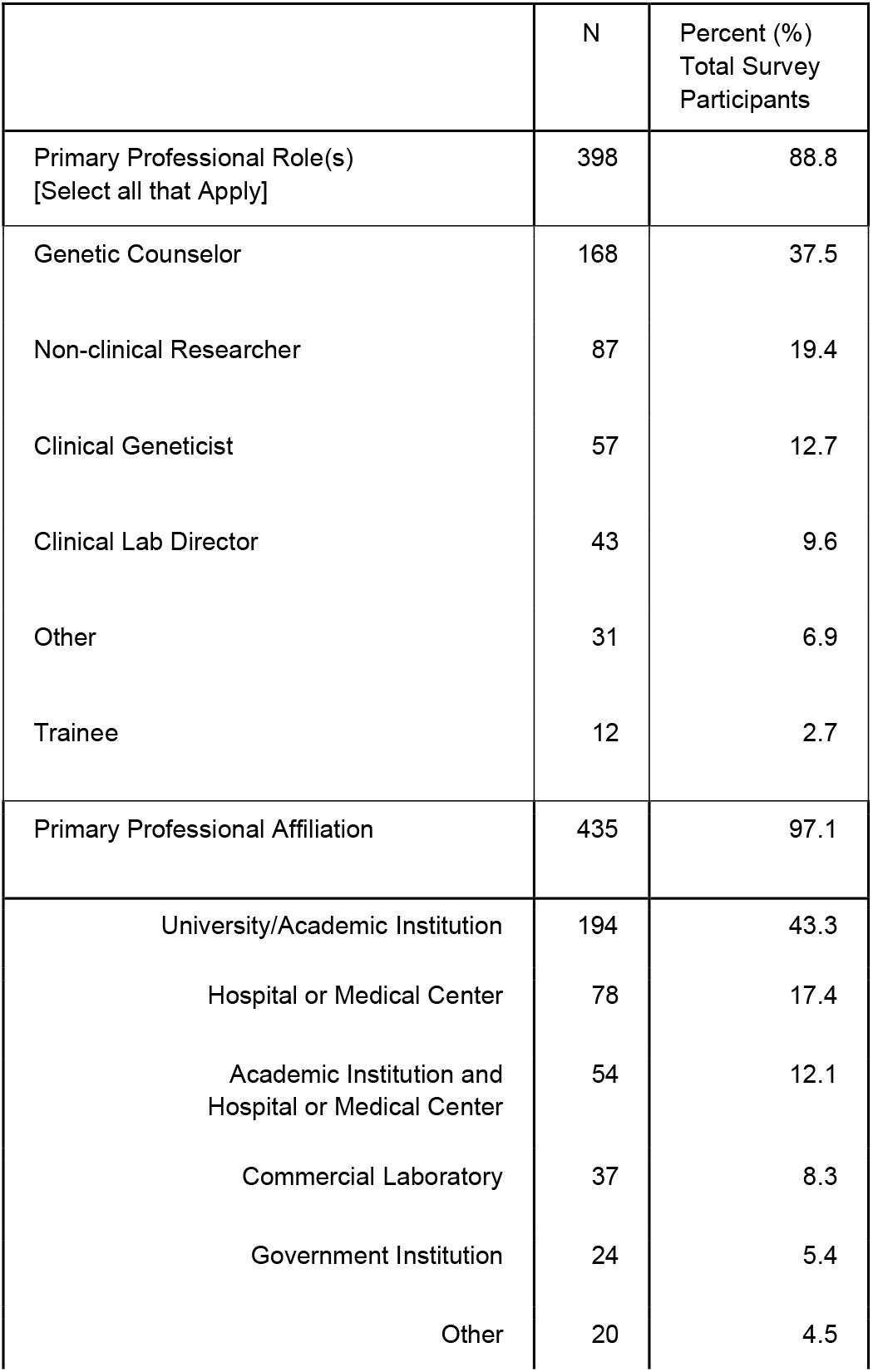
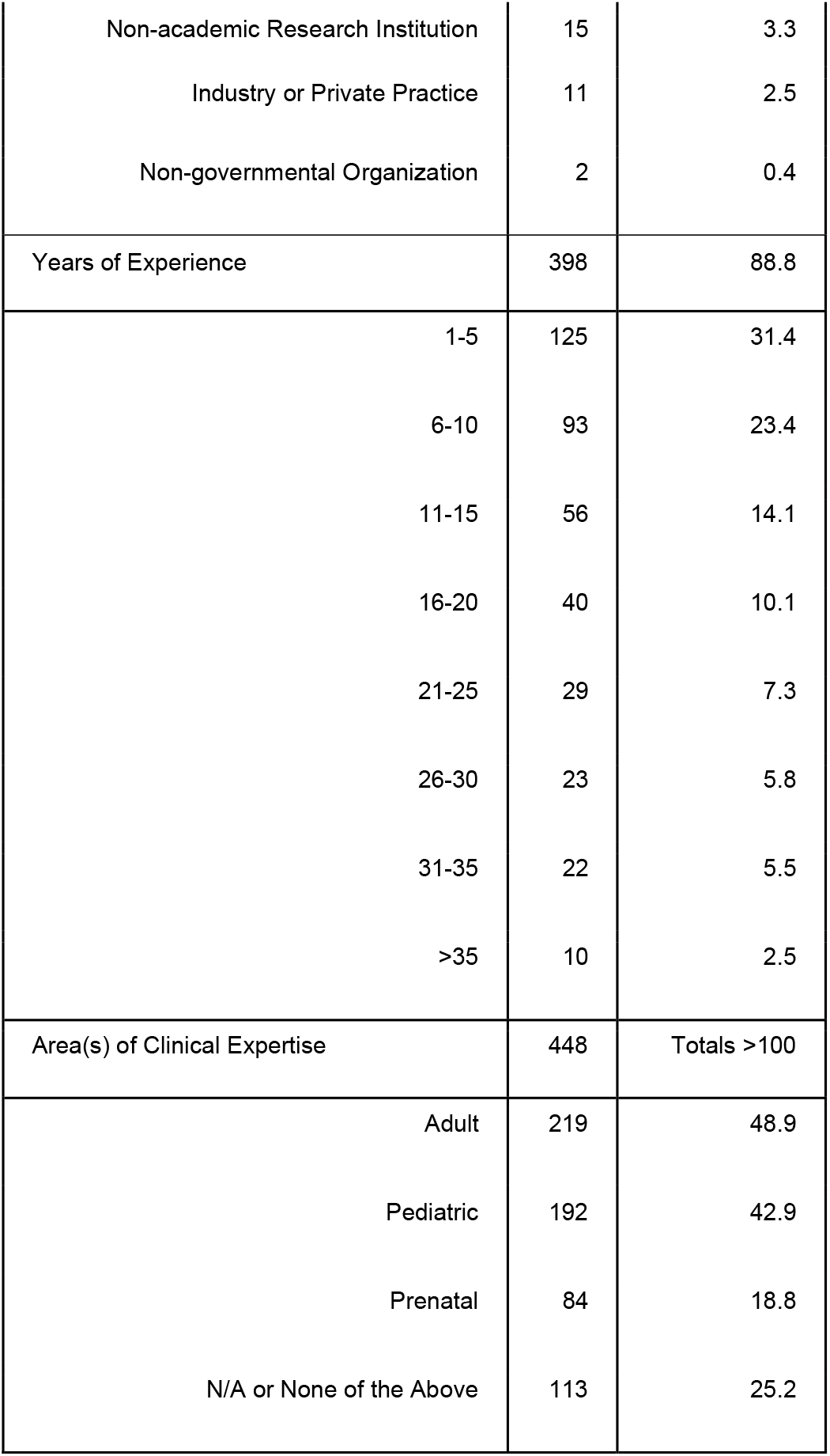
Survey Participant Professional Roles and Affiliations. Most (but not all) survey participants provided information about their professional roles, affiliations, years of experience, and clinical area(s) of expertise. In some cases, total percentages add up to >100% due to participants selecting more than one response. Total percentages that are <100% indicate questions to which not all survey participants responded.

|  | N | Percent (%)<br>Total Survey<br>Participants |
| --- | --- | --- |
| Primary Professional Role(s)<br>[Select all that Apply] | 398 | 88.8 |
| Genetic Counselor | 168 | 37.5 |
| Non-clinical Researcher | 87 | 19.4 |
| Clinical Geneticist | 57 | 12.7 |
| Clinical Lab Director | 43 | 9.6 |
| Other | 31 | 6.9 |
| Trainee | 12 | 2.7 |
| Primary Professional Affiliation | 435 | 97.1 |
| University/Academic Institution | 194 | 43.3 |
| Hospital or Medical Center | 78 | 17.4 |
| Academic Institution and<br>Hospital or Medical Center | 54 | 12.1 |
| Commercial Laboratory | 37 | 8.3 |
| Government Institution | 24 | 5.4 |
| Other | 20 | 4.5 |
| Non-academic Research Institution | 15 | 3.3 |
| Industry or Private Practice | 11 | 2.5 |
| Non-governmental Organization | 2 | 0.4 |
| Years of Experience | 398 | 88.8 |
| 1-5 | 125 | 31.4 |
| 6-10 | 93 | 23.4 |
| 11-15 | 56 | 14.1 |
| 16-20 | 40 | 10.1 |
| 21-25 | 29 | 7.3 |
| 26-30 | 23 | 5.8 |
| 31-35 | 22 | 5.5 |
| >35 | 10 | 2.5 |
| Area(s) of Clinical Expertise | 448 | Totals >100 |
| Adult | 219 | 48.9 |
| Pediatric | 192 | 42.9 |
| Prenatal | 84 | 18.8 |
| N/A or None of the Above | 113 | 25.2 |

**Table 2.** Participant Self-reported Sex, Gender, Race, and Ethnicity. All survey respondents were asked to write free-text responses describing their identities with regard to sex, gender, race, ethnicity, and ancestry. They were also asked to select their race and ethnicity from multiple-choice options. Shown here are aggregate results of free-text responses about sex and gender, as well as aggregate responses to the multiple-choice race and ethnicity question.

|  | N | Percent (%) Total Survey Participants |
| --- | --- | --- |
| Sex and Gender (Free Text Response) | 448 | 100 |
| Female | 281 | 62.7 |
| Male | 98 | 21.9 |
| Transgender | 1 | 0.22 |
| Irrelevant (not related to sex or gender) or Missing Response | 68 | 15.2 |
| Race and Ethnicity (Multiple Choice) | 448 | 100 |
| *Single Selection* | 373 | 83.3 |
| American Indian/Alaska Native | 0 | 0.0 |
| Asian | 43 | 9.6 |
| Black or African American | 7 | 1.6 |
| Hispanic or Latino | 6 | 1.3 |
| Native Hawaiian or Other Pacific Islander | 0 | 0.0 |
| White | 317 | 70.8 |
| *Multiple Selections* | 14 | 3.1 |
| American Indian/Alaska Native and White | 3 | 0.7 |
| American Indian/Alaska Native, Hispanic or Latino, and White | 3 | 0.7 |
| Asian and White | 4 | 0.9 |
| Asian and Hispanic or Latino | 1 | 0.22 |
| Black or African American and White | 1 | 0.22 |
| Hispanic or Latino and White | 1 | 0.22 |
| All of the Above | 1 | 0.22 |
| *Missing Response (Race and Ethnicity)* | *61* | *13.6* |

Sixty percent of survey participants report interpreting or curating genetic variants for a variety of purposes (N=271), including for research (N=170, 63%), providing clinical genetic testing reports to healthcare providers (N=125, 46%), informing the diagnosis of a patient with a genetic disorder (N=122, 45%), reclassifying variants and/or verifying laboratory results (N=121, 45%), preparing for a discussion with a patient (N=120, 44%), and curating genetic variants for large-scale consortium efforts, such as ClinGen (N=99, 36%).

Among those who provided information about their geographic location (N=399, 89% of total participants) 93% are in North America, with 347 in the U.S. and 23 in Canada. Outside North America, 19 participants are from Europe, and the remaining individuals are from Australia, Japan, Mexico, Oman, Singapore, and South Korea. The majority of respondents self-identified as white (71%), and 63% identified as female.

### Adjusted Response Rate

The overall response rate (4.2%) was adjusted for overlap in professional society membership and consortium affiliations. Respondents were asked to indicate all relevant professional affiliations with the organizations sampled, which facilitated the elimination of double counting such that each survey respondent was counted only once in the denominator of all possible respondents. Table S1 indicates response rates that are specific to each disseminating organization, which are also individually adjusted to ensure each study participant was counted in only one targeted recruitment group. Due to residual overlap in organizational membership among survey *non*-responders, our response rate is likely underestimated. Furthermore, with a snowball approach (employed during the latter portion of the recruitment period), the number of potential respondents cannot be estimated, so these responses are not counted in the overall response rate for the survey.

### Collecting REA Data in Practice

Nearly all participants (N=422, 94%) collect or use information about “population identity, e.g., population allele frequencies, self-reported race or ethnicity, and/or ancestral origins” in their work. The majority of participants reported using this information for the purpose of clinical practice alone (N=178, 39%) or for the purposes of both clinical practice and research (N=130, 29%). Among respondents who see patients, order genetic tests, and/or communicate results (N=268), only 5 individuals (2%) report that no REA information is used in their work, and 4 (2%) were not sure. Race and/or ethnicity is most often obtained directly from the patient (N=204, 94%), or from the patient” s medical record (N=87, 40%). Some reported that race and/or ethnicity is recorded by another care provider, “possibly without verifying directly with the patient” (N=39, 18%).

### Conceptualizing REA in Clinical Genetics

Figure 1 illustrates differences among survey respondents in their agreement with select phrases offered to describe the terms “race”, “ethnicity”, and “ancestry”. Nearly two-thirds of survey participants indicated that they felt “ancestry” was well described by the term “genetic lineage group” (N=295, 66%). “Cultural group” was perceived to describe “ethnicity” well or very well by 46% of participants (N=205). No term or phrase was considered a good description of “race”, and ∼80% of participants felt that none of the three REA terms were well described by “species group”, “lifestyle/behavioral group”, or “religious group”. Beyond these areas of relative agreement, there was otherwise substantial heterogeneity in respondents’ agreement with the definitions provided.

**Figure 1.**
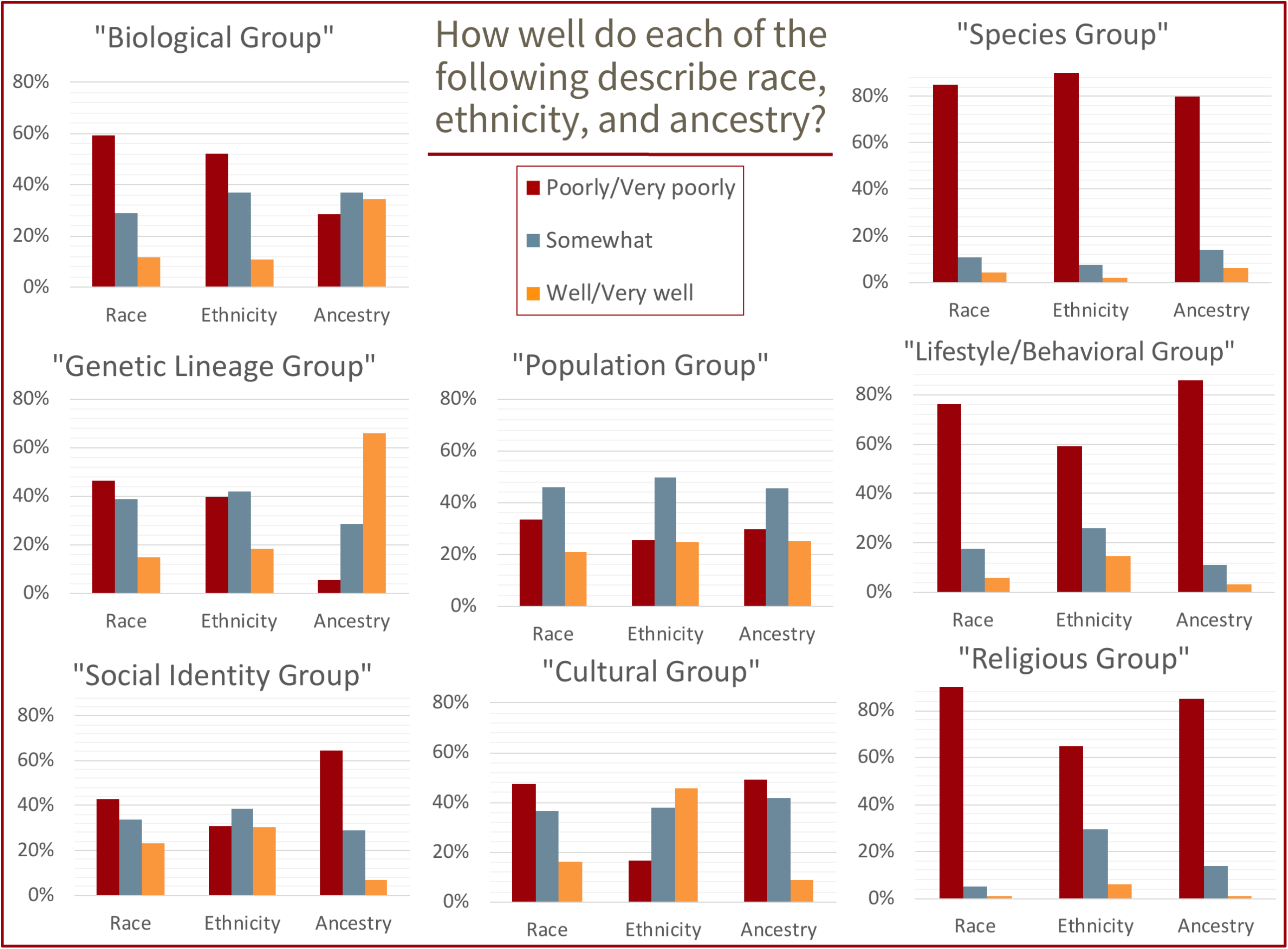
Perceived Definitions of Race, Ethnicity, and Ancestry. Each cluster of bars corresponds to a complete set of survey responses, such that 100% of the participants replied that each description fit each term (“race”, “ethnicity” or “ancestry”) either very well, well, somewhat, poorly, or very poorly.

Figure 2 illustrates respondent confidence in describing differences among “race”, “ethnicity”, and “ancestry”, in clinical genetics and in general. More than two-thirds of participants reported being “somewhat” or “not at all confident” in their ability to distinguish among REA terms, both in general and as they relate to genetics and clinical care. However, there was a marked shift in the proportion of survey participants who responded that they are “confident” when asked about their ability to distinguish among terms in the context of genetics and clinical care. Roughly twice as many participants report being “not at all confident” about the terms in general (N=123, 27%), compared with those who felt “not at all confident” about the terms related to their line of work in genomics and clinical care (N=62, 14%).

**Figure 2.**
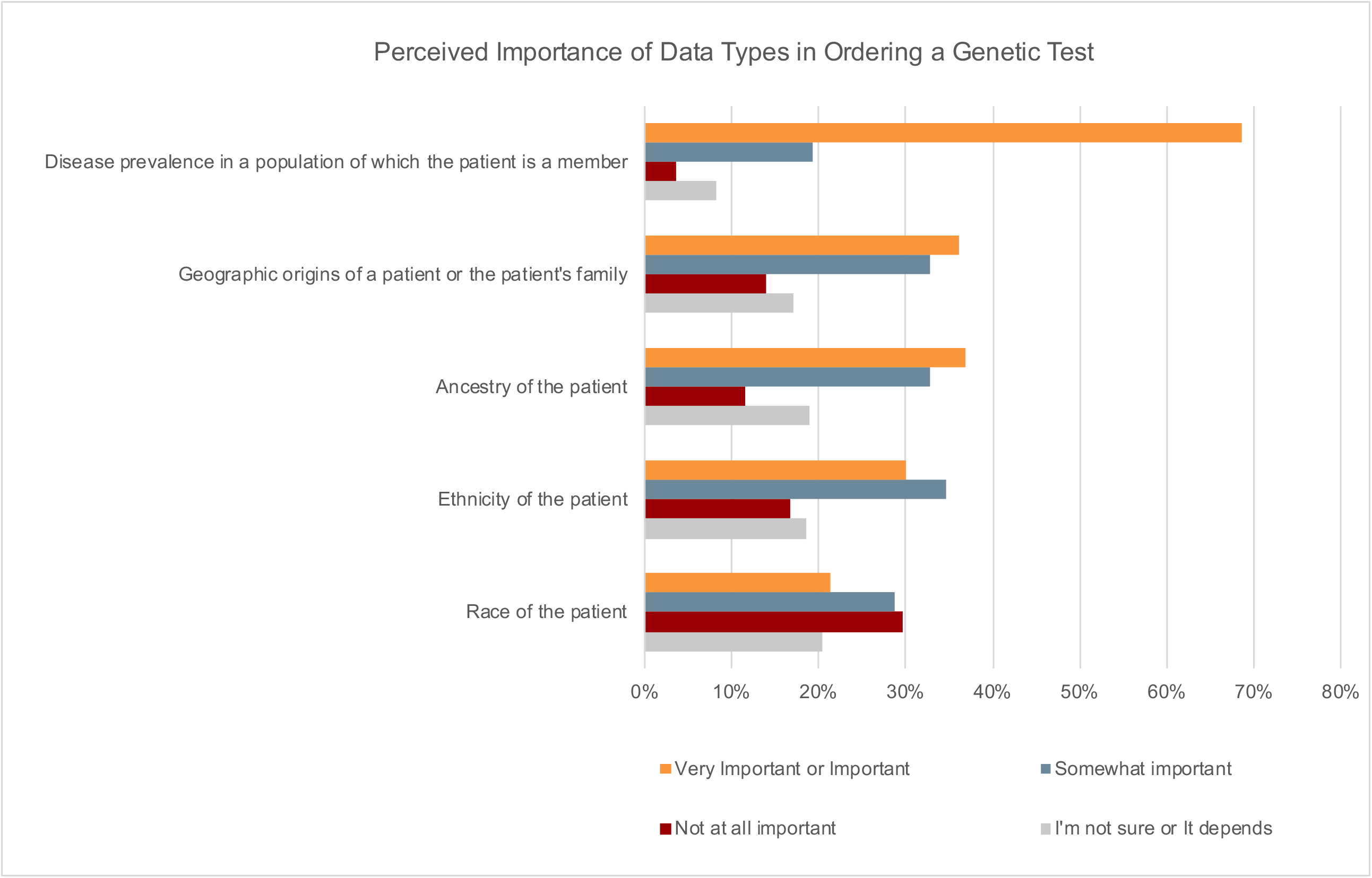
Perceived Importance of Patient Data Types in Ordering a Genetic Test. Clinical professionals (N=268) indicate the degree of importance for each type of information (disease prevalence in a population, geographic origins and REA of a patient) for the purpose of ordering a genetic test.

### Use of REA in Clinical Genetics

Two hundred sixty-eight (60%) of survey participants indicated that they order clinical genetic tests and/or return results to patients in their own work and answered a series of questions about patient-facing activities, including perceptions about the relevance of REA for clinical decision-making. Patient race (21%), ethnicity (30%), ancestry (37%), geographic origins of a patient or their family (36%), and disease prevalence in a population of which the patient is a member (69%) were all considered important or very important information for the purpose of ordering a genetic test (Fig.3). When asked about other types of clinical decision-making, 49% percent of participants agreed that REA would be relevant for obtaining consent.

Clinical professionals were also asked about clinical factors and circumstances that would likely motivate them to discuss REA with a patient. These included whether test results were positive (48%), revealed a variant of uncertain significance (VUS) (50%), or whether the patient was from a racial or ethnic minority group (53%) (Table 5). A large majority (N=189, 90%) agreed that REA may be relevant for contextualizing genetic test results for patients. Across all clinical scenarios, there was substantial heterogeneity in the perceived relevance of REA, and for most categories up to a third responded “I’m not sure.”

### REA in Genetic Variant Interpretation

When asked about the importance of REA for clinical interpretation of genetic variants, respondents perceived these measures as highly important, though practical use and implementation was reportedly much lower. Table 3 shows the degree to which participants reported each of these measures to be important. Some participants pointed out that the type of information most likely to inform interpretation depends on the information that is available. Specifically, ancestry was considered more relevant than race or ancestry for clinical interpretation but was reportedly less often available and used in practice.

**Table 3.** Importance of Race, Ethnicity, and Ancestry in Clinical Variant Interpretation. Survey respondents who reported having a professional role in clinical care, such as seeing patients and ordering genetic tests, were asked to evaluate the importance of race, ethnicity, and ancestry for the purpose of clinical variant interpretation. Results are shown here by the multiple-choice (Likert scale) options provided.

| N=271 | RACE | ETHNICITY | ANCESTRY |
| --- | --- | --- | --- |
| Very Important | 22 (8.1%) | 22 (8.1%) | 43 (15.9%) |
| Important | 48 (17.7%) | 67 (24.7%) | 81 (29.9%) |
| Somewhat important | 85 (31.4%) | 70 (25.8%) | 71 (26.2%) |
| It depends | 54 (19.9%) | 58 (21.4%) | 49 (18.1%) |
| Not at all important | 36 (13.3%) | 29 (10.7%) | 10 (3.7%) |
| I'm not sure | 26 (9.6%) | 25 (9.2%) | 17 (6.3%) |

Fewer than half of participants reported that any of the measures of race, ethnicity, geographic origin(s), or ancestry were likely to inform clinical variant interpretation in practice. For example, despite ancestry being reported as most consistent with the concept of genetic lineage or inheritance and as the most important to clinical variant interpretation (46% responding that it was important or very important; Table 3), the majority of survey participants (85%) still report that genetic ancestry is rarely (N=31, 12%) or never (N=196, 73%) calculated from a patient’s DNA for the purpose of clinical variant interpretation (See Appendix for methods that are used by respondents to calculate ancestry). Participants also report that clinical lab reports either rarely (N=40, 18%) or never (N=161, 74%) contain ancestry estimates based on genetic data when reporting carrier or diagnostic test results.

The ACMG/AMP Guidelines for the Interpretation of Sequence Variants (Richards et al., 2015) call for the use of population databases, and rely on allele frequency data to guide the assessment of genetic variants. Table 4 shows the proportion of participants who report using each type of REA data to inform clinical variant interpretation. Consistent with the ACMG/AMP recommendations, both population allele frequencies (86%) and the related concept of absence of a variant from “population databases” (83%) are most widely considered informative for interpretation and the majority of participants indicated using multiple publicly-available databases to obtain this information.

**Table 4.** Data Types Most Likely to Inform Clinical Variant Interpretation. A subset of clinical genetics professional survey respondents (N=271) selected the type(s) of data they are “most likely” to use when conducting clinical variant interpretation.

| Data type | Individuals (N) who reported whether they are “most likely” to use each type of data to inform variant interpretation. |
| --- | --- |
| Population allele frequencies | 233 (86%) |
| Whether a variant has been seen before in a population database | 224 (82.7%) |
| Ancestry of the patient or population in which the variant was observed | 120 (44.3%) |
| Geographic origin(s) of the patient’s family or population in which the variant was observed | 108 (40%) |
| Ethnicity of the patient or population in which the variant was observed | 103 (38%) |
| Race of the patient or population in which the variant was observed | 89 (32.8%) |
| All of the Above | 49 (18.1%) |

**Table 5.**
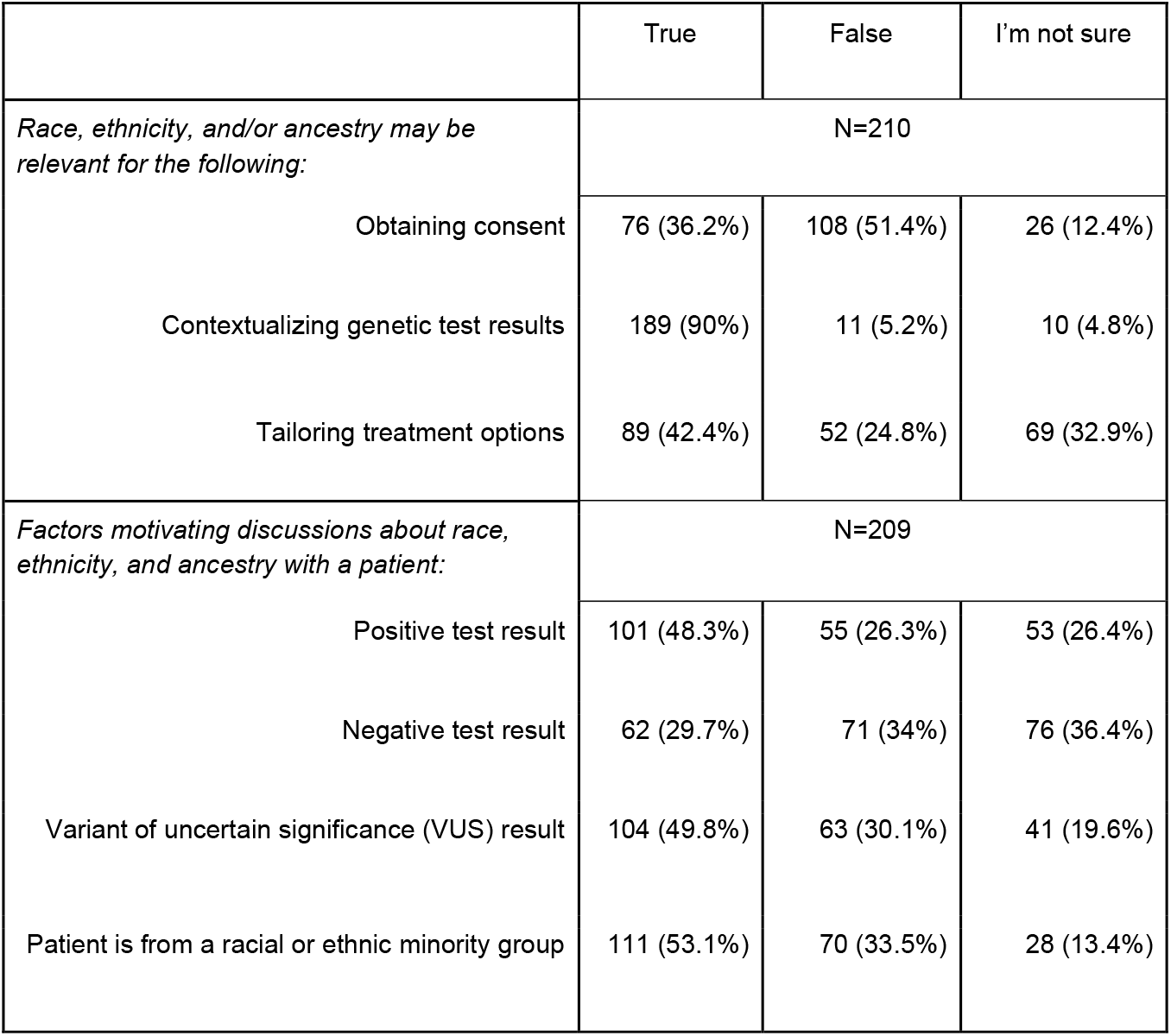
Perceived Importance of Discussing Race, Ethnicity, and Ancestry with Patients Undergoing Clinical Genetic Testing. A subset of survey respondents who see patients responded to true/false questions about the type of clinical functions for which race, ethnicity, or ancestry may be relevant and factors that might motivate them to discuss these with a patient.

### Necessity of REA Guidelines for Clinical Genetics

The majority of respondents who answered questions on the need for new guidelines (N=425) felt that guidelines would be helpful for the collection and use of REA in clinical genetics (N=276, 65%). Thirteen participants (3%) disagreed and offered reasons as to why they did not feel new guidelines would be helpful. Common themes include the complexity of issues that cannot be easily digested or distilled, that guidelines would be ineffective or burdensome to implement, and that issues of diversity measures are irrelevant to the field.

## Discussion and Recommendations

This study illuminates clinical genetics professionals’ beliefs about the meaning and utility of REA, as well as its reported use in clinical practice. While most respondents considered REA at least somewhat important for clinical interpretation and ordering or communicating the results of genetic tests, there were inconsistent understandings of these concepts, and differences of opinion on how these terms should be used in clinical genetics, and for what purposes. Interestingly, there appeared to be a disconnect between the type of information that is considered most useful by clinical genetics professionals and the type of information they reported having access to (and using) in practice. Highly variable opinions, definitions and perceived utility of REA for clinical interpretation and the delivery of genomic medicine is consistent with the established view that definitions of “race” and “ethnicity” are cultural, dynamic, and change over time (Smedley & Smedley, 2012; Omi & Winant, 2014; Roth, 2016). These results support the need to standardize the collection and use of REA in clinical genetics.

### Perceived Definitions and Utility of REA Vary Widely

Our results indicate that participants are most likely to understand ancestry in biologic terms (as ‘genetic lineage’ groups) and that in the context of clinical genetics they evaluate the importance of ancestry as higher than that of race or ethnicity. However, this measure is the least available and infrequently used in practice. Likely due to its lack of availability in practice, survey respondents did not express a greater reliance on this measure for clinical variant interpretation than on race or ethnicity. Nearly 70% of participants said that disease prevalence in a population of which a patient is a member is important for ordering genetic tests, but without access to genetic ancestry, clinicians must rely on self-reported (or assigned) measures of race and ethnicity in determining to which population a patient should be attributed. Respondents confirm this information is most often obtained either directly from a patient or an existing medical record and used as a proxy for genetic ancestry. However, as shown previously (Popejoy et al., 2018), there is also a lack of consistency in how such self-reported measures are collected on clinical laboratory requisition forms, if at all, and some laboratory directors reported feeling that this information is of limited usefulness. This is particularly problematic for individuals with diverse or multiple ancestral backgrounds, as a single ancestral population cannot be assigned even with the ability to calculate or obtain genetic ancestry estimates. A similar challenge exists in the context of clinical variant interpretation, given that the vast majority of participants use population allele frequencies to determine whether a variant is rare and thus considered more likely to be pathogenic. Race and ethnicity are socio-cultural (not biological) in nature, so while they may be useful as a proxy for ancestral background in some cases, this approach is not recommended. In order for this assessment to be relevant, greater representation of diverse populations in allele frequency databases is essential.

There was notable disagreement and uncertainty among survey participants with regard to the clinical scenarios that would be most likely to motivate the use of REA in clinical decision-making or discussions with patients. Most survey participants expressed that REA are each at least *somewhat* important for ordering genetic tests, interpreting results, and communicating with patients. However, participants do not agree about the factors that would contribute to the relevance of REA, which indicates that they may in fact have less practical utility than commonly assumed. It may also suggest that the use of REA is highly contextual such that it becomes important for ordering tests, billing, and medical coverage in some cases and in others it can inform discussions about the limitations of testing in certain populations.

There is evidence to suggest that descriptions of patient race and ethnicity in electronic medical records (EMRs) are often inaccurate (Klinger et al., 2015), which calls into question the utility of this information. Because race, ethnicity, and ancestry each convey and represent a distinct type of information, such as one’s physical or social environment, lived experiences (e.g., racism), cultural traditions (e.g., nutrition, lifestyle), and genomic background, it is not recommended to collapse these measures or use one as a proxy for another. In order to mitigate the continued use of REA in clinical genetics without rigorous scientific evaluation and standardization of this practice, we recommend that applications of this information should be justified through formal inquiry on the basis of effectiveness and necessity. It may be that using REA is not appropriate in all clinical scenarios, or that each measure may have specific, unique utility for particular applications. If REA is necessary and effective for the delivery of health care in certain clinical settings, formal and consistent definitions must be developed through a deeply interdisciplinary and deliberative process, then widely disseminated and adopted across clinical laboratories. Future efforts to determine what information about patients can and should be required will need to combine stakeholder engagement, policy analysis, and research on the utility of these measures in specific clinical settings.

### Interpreting Variants from Diverse Populations

Respondents reported high perceived utility of REA information in the context of variant interpretation. Nevertheless, there is little guidance regarding how such information should be used in clinical variant interpretation and curation. The ACMG/AMP Guidelines for variant interpretation suggest that absence of a variant from “population databases” is moderate evidence for the pathogenicity of a variant (PM2). However, most databases that clinicians and researchers use to verify the presence or absence of a variant are not representative of the global diversity in human genomic variation (Lek et al., 2016; Landry et al., 2018). Further complicating the matter, self-reported race or ethnicity is often used as a proxy for genetic ancestry (since the latter is rarely available), so determining which genomic background or ancestral population is most relevant for the assessment of allele frequencies and the validity of PM2 for a particular patient remains a challenge. Future discussions among interdisciplinary researchers could focus on solutions to this particular challenge as it has a direct impact on the reported pathogenicity of variants.

### Study Limitations

Although the results of this survey provide important insights for understanding how REA are used in clinical genetics, there are limitations. Our survey participants were heavily skewed toward individuals who are involved in U.S.-based research consortia ClinGen and CSER. Additionally, our method of dissemination and recruitment may have biased our sample to individuals who are already interested in the topic of diversity in genetics, and thus may think more deeply about the role of these attributes in both research and clinical care. Due to overlap in organizational membership among survey non-responders (which could not be accounted for), the response rate is underestimated. We may have also underestimated the proportion of clinical professionals who make their own determination about a patient’s race or ethnicity, because those who report obtaining this information from a medical record were not asked how this information was placed in the medical record. Finally, while the lack of racial and ethnic diversity among our study participants may appear to be a limitation, it appears reflective of the demographics of the clinical genetics professions and is likely a representative sample of our target study population. It may be useful in future studies to over-sample a more diverse sample of clinical genetics professionals, in order to shed light on how perspectives may differ.

### Conclusions and Next Steps

The results of this survey establish a baseline understanding of how REA are perceived and utilized by clinical genetics professionals, and our results show that there is little standardization or consistency. As a result of considerable individual-level variability in beliefs about how REA should inform clinical genetics and a lack of generally accepted understanding or standards, there is ample opportunity for bias to influence the research and implementation of precision medicine. Therefore, we recommend further research and suggest that REA utilization in clinical genetics be standardized, evidence-based, and justified, in order for the implementation of genomic medicine to be consistent, scientifically valid, and ethically responsible.

The National Academy of Science, Engineering and Medicine (NASEM) standards for the development of clinical practice guidelines recommend a systematic review of evidence from the literature about a given practice or protocol, and the subsequent assemblage of a multi-disciplinary group of experts to serve as a guideline-development committee (Institute of Medicine, 2011). Therefore, we recommend that diverse stakeholders and representatives from patient populations, clinical laboratories, educational and research institutions, funding agencies, professional organizations, accreditation bodies, scientific journals, and multi-institutional research consortia collaborate to develop a set of standards and recommendations for the clinical genetics community about the utility and use of REA to inform precision medicine.

## Data Availability

Data may be available upon request to the corresponding author.

## Appendix

### Supplemental methods

#### Cognitive Interviews to Validate Survey Instrument

In order to refine survey questions for clarity and understanding by the intended respondents, cognitive interviews were conducted by a single interviewer (A.B.P.) with 11 representatives from target participant groups including non-clinical genetics researchers and clinical genetics professionals (i.e., clinical geneticists, clinical genetics laboratory directors and genetic counselors), most of whom were recruited through CSER. During cognitive interviews, participants were asked to read survey questions aloud and talk through their understanding and interpretation of each question. Their ability to determine or recall the requested information was observed. If a participant had difficulty or was unable to answer a question, the interviewer discussed the question further with the participant to assess interpretation issues and provided alternative phrasing until the intended question was clearly comprehended. As interview participants read and answered survey questions while speaking aloud their thought process, the interviewer asked follow-up questions about their interpretation of the questions to ensure consistency with the intended design.

Audio and video were recorded for online interviews (N=8), and audio was recorded for in-person interviews (N=3). Every three interviews, the interviewer used notes and recordings to revise survey questions to reflect language that was best understood and interpreted by the majority of participants. As the language and format of questions was refined through cognitive interviews, participants were able to understand more questions without needing clarification, and the ability of participants in later interviews to correctly interpret and answer questions was improved relative to earlier ones. The revised survey was reviewed and approved by ClinGen ADWG members and CSER investigators.

#### Survey Questions (All respondents)

1. Do you collect or use any information about population identity (E.g., population allele frequencies, self-reported race or ethnicity, and/or ancestral origin(s) of a patient or family members) in any aspect of your research or clinical practice? (Yes/No)
2. In your opinion, and based on your own understanding, how well does the term “biological group” describe each of the following? (Required) – [Race/Ethnicity/Ancestry]
3. In your opinion, and based on your own understanding, how well does the term “cultural group” describe each of the following? (Required) – [Race/Ethnicity/Ancestry]
4. In your opinion, and based on your own understanding, how well does the term “genetic lineage group” describe each of the following? (Required) – [Race/Ethnicity/Ancestry]
5. In your opinion, and based on your own understanding, how well does the term “lifestyle/behavioral group” describe each of the following? (Required) – [Race/Ethnicity/Ancestry]
6. In your opinion, and based on your own understanding, how well does the term “population group” describe each of the following? (Required) – [Race/Ethnicity/Ancestry]
7. In your opinion, and based on your own understanding, how well does the term “religious group” describe each of the following? (Required) – [Race/Ethnicity/Ancestry]
8. In your opinion, and based on your own understanding, how well does the term “social identity group” describe each of the following? (Required) – [Race/Ethnicity/Ancestry]
9. In your opinion, and based on your own understanding, how well does the term “species group” describe each of the following? (Required) – [Race/Ethnicity/Ancestry]
10. How often do you interpret or curate genetic variants in either a clinical or research setting? (Multiple choice)

<u>The following questions were asked only to those who reportedly conduct variant interpretation</u>:

11. How often do you order genetic tests or return results to patients in a clinical care setting? (Multiple choice)
12. For what purpose(s) do you typically interpret genetic variants? (Select all that apply)
13. When interpreting genetic variants, which of the following kind(s) of data are most likely to inform your interpretation? (Select all that apply / Free text option)
14. If information about race, ethnicity, and/or ancestry is used in clinical variant interpretation, from which of the following source(s) are these data most likely obtained? (Select all that apply / Free text option)
15. Which of the following resource(s) do you use frequently in the curation of genetic variants? (Select all that apply / Free text option)
16. Which of the following resources are you likely to use when applying population-level data to variant curation or interpretation? (Select all that apply / Free text option)
17. To your knowledge, what information about population-level data is typically included in reports from clinical testing laboratories? (Select all that apply)
18. How important are the following in the clinical interpretation of genetic variants? – [Race/Ethnicity/Ancestry]
  a. Since you indicated that “it depends” how important race, ethnicity, and/or ancestry are for clinical variant interpretation, please feel free to elaborate here. (Free text response)
19. In your opinion, what is the relevance of race, ethnicity, and/or ancestry information in the context of clinical variant curation or interpretation? (Multiple choice)
20. How often do you calculate genetic ancestry from a patient” s DNA for the purpose of clinical interpretation? (Multiple choice)
  a. When genetic ancestry is calculated by you and/or your colleagues for the purpose of clinical interpretation, what method(s) are most often used?
  b. Which reference population panel(s) do you and/or your colleagues use when estimating genetic ancestry for clinical interpretation?

<u>The following questions were asked only to those who reportedly work in genomic medicine and clinical care, including ordering and interpreting genetic tests, reporting results to patients, etc.</u>:

21. How often do you receive estimates of a patient” s genetic ancestry (calculated from DNA) in addition to carrier or diagnostic test results on a clinical lab report? (Multiple choice)
22. If information about a patients’ race, ethnicity, or ancestry is used in any aspect of your work as a clinician, from where are these data likely obtained? (Select all that apply / Free text response)
23. For the purposes of ordering a genetic test, how important do you consider each of the following type(s) of data? (Likert scale of importance)
  a. Race of the patient
  b. Ethnicity of the patient
  c. Ancestry of the patient
  d. Geographic Origin(s) of the patient
  e. Disease prevalence in a of which the patient is a member
    i. Since you indicated that “it depends” how important certain identity or other measures are for ordering a genetic test, please feel free to explain here. (Free text response)
24. In your opinion, what is the relevance of race, ethnicity, and/or ancestry information in decision-making about ordering a genetic test? (Free text response)
25. Race, ethnicity, and/or ancestry may be relevant for obtaining consent from patients for a clinical diagnostic genetic test. (True/False)
26. Race, ethnicity, and/or ancestry may be relevant for contextualizing genetic test results in discussions with a patient. (True/False)
27. Race, ethnicity, and/or ancestry may be relevant for tailoring treatment options to patients. (True/False)
28. A positive test result would likely motivate me to discuss race, ethnicity, or ancestry (REA) for relevant traits and conditions. (True/False)
29. A negative test result would likely motivate me to discuss REA for relevant traits and conditions. (True/False)
30. If the patient has a variant of uncertain significance (VUS) in a gene of interest, I am more likely to discuss REA with them. (True/False)
31. If the patient is a member of an ethnic or racial minority population, I am more likely to discuss REA with them. (True/False)
32. Are there other factors (beyond those listed in the previous questions) that might motivate you to discuss race, ethnicity, and/or ancestry with a patient? (Yes/No)
  a. a. Since you indicated that there are other factors that might motivate you to discuss race, ethnicity, and/or ancestry with a patient, please list or describe them here.
33. In your opinion, are there particular traits or conditions for which you think it most important to consider race, ethnicity, and/or ancestry? (Yes/No)
34. How confident are you in describing the similarities and differences among race, ethnicity, and ancestry? (Multiple choice)
35. How confident are you in discussing how race, ethnicity, and ancestry are related to genomics and clinical care? (Multiple choice)

<u>The following questions were asked to all respondents:</u>

36. Do you believe that new guidelines would be helpful to guide the collection, use, and/or communication of race, ethnicity, and ancestry information in clinical genomics? (Multiple choice)
  a. a. Please feel free to elaborate on why you indicated “other” in response to the previous question about whether new guidelines would be helpful to guide the collection, use, and/or communication of race, ethnicity, and ancestry information in clinical genomics. (Free text response)
  b. b. For which of the following purposes do you agree that it would it be useful to develop standards or guidelines in clinical genomics? (Select all / Free text)
  c. c. Please feel free to elaborate on how standards or guidelines about the use of race, ethnicity, and/or ancestry would be helpful for researchers and professionals working in clinical genomics. We welcome any suggestions or comments.
  d. d. Why do you believe that no additional guidelines are needed for the use of race, ethnicity, and or ancestry information? (Select all that apply / Free text response)
  e. e. Since you indicated “other” reasons that there is no need for additional guidelines for the use of race, ethnicity, and/or ancestry information, please feel free to elaborate here.
37. Which of the following organizations or consortia are you a member of, or affiliated with? (Select all that apply)
38. Which of the following entities best represents your employment affiliation(s)? (Select all that apply / Free text response)
39. Please indicate the country in which your primary employer is located. (Multiple choice)
  a. a. Please indicate in which U.S. state your primary employer is located.
40. Please specify your role(s) with regard to clinical genomics. (Select all that apply / Free text response)
41. For how many years have you been working in this field?
  a. <u>Clinical professionals only</u>: Do you work primarily in Prenatal, Pediatric or Adult Medicine? (Select all that apply)
42. How do you personally identify on the basis of each of the following? (Fill in the blank with your own words to describe your identity.)
  a. Sex
  b. Gender
  c. Race
  d. Ethnicity
  e. Ancestry
43. The following choices are designated by the U.S. Office of Management and Budget (OMB) as the official racial categories used for reporting demographics in the federal government. Please check the box(es) that best correspond(s) to your personal identity.
44. Which of the two previous questions about your own personal identity would you prefer to be asked in the future?
  a. Why did you prefer to answer the open-ended identity question?
  b. Why did you prefer to answer the multiple-choice identity question?
45. Have you seen any version of these survey questions before (E.g., as part of the development or validation of the survey)? (Yes/No/I” m not sure/Free text response)

#### Supplemental Results

##### Ancestry Analysis for Clinical Variant Interpretation

Among those who report doing *any* ancestry analysis for clinical variant interpretation (N=60), the most common methodological approaches are Principal Component Analysis (PCA, N= 34, 56%) and admixture analysis (N= 16, 26%). Five participants (8%) report using multidimensional scaling, while another 15 (25%) do not know what method is used to calculate ancestry. For relevant methods, the most commonly used reference panels for ancestry inference are 1000 Genomes (N= 30, 73%), HapMap (N=10, 24%), and Human Genome Diversity Project (HGDP, N=11, 27%) or some combination thereof; some also report using internal reference panels either alone or in combination with others (N=11, 27%).

#### Supplemental Data

Table S1. Survey Dissemination and Response Rates.

## Acknowledgments

Funding for this study was provided by the National Human Genome Research Institute (NHGRI) Clinical Genome Resource (ClinGen): 5U41HG009649-03; the Clinical Sequencing Evidence-generating Research (CSER) Coordinating Center: U24HG007307; the UCSF Program in Prenatal and Pediatric Genome Sequencing (P3EGS): U01HG009599; and a Chan-Zuckerberg Investigator Award (J.Y.Z.).

Individuals who contributed to the development of this study through working group discussions and/or participation in refinement of the survey instrument include Vence Bonham, Eimear Kenney, Manuel Rivas, Jonathan Berg, Tim Thornton, Laura Amendola, Kate Foreman, Gail Jarvik, Mary Norton, Sarah Scollon, Jacqueline Odgis, and other anonymous contributors.

Dissemination of the survey was supported by authors and the following individuals, on behalf of the affiliated organizations and consortia: Jeffrey Ou (CSER); Danielle Azzariti (ClinGen); Candice Miller (ABGC); Denise Calvert (ACMG); Mona Miller (ASHG). Maria Cerezo (GWAS Catalog) and others contributed to targeted social media posts. Lauren Hicks and Catherine Gooch contributed a manually curated email list of publicly listed clinical geneticists.

The authors appreciate feedback on earlier versions of this paper by attendees to the Stanford Center for Biomedical Ethics writing seminar.

## Web Resources

U.S. Census Bureau: https://www.census.gov/topics/population/race/about.html

